# Neural Network-Based Assessment of Coronary Angiogram Flow

**DOI:** 10.1101/2024.12.03.24318433

**Authors:** Jesse M. Resnick, Ismael Z. Assi, Aishwarya Pastapur, Ajit Mullasari Sankardas, C. Alberto Figueroa, Brahmajee K. Nallamothu

## Abstract

Adequate blood flow through the coronary tree is critical for maintaining cardiac perfusion. Coronary angiography has the potential to provide rich and dynamic hemodynamic information, however, current strategies to assess flow through the coronary vessels depend on either subjective expert opinion (TIMI flow grade) or laborious frame-by-frame anatomical analysis (TIMI frame count and quantitative flow ratio). Here we present a strategy for automated characterization of bulk flow through the coronary tree using the right coronary artery as an example. We leverage the AngioNet neural network to generate sequential segmentations of angiograms, create time series of the summed segmentation intensities (i.e., a contrast intensity profile), and quantitatively characterize the filling and washout phases of these intensity profiles. We demonstrate that AngioNet-derived frame counts and normalized mean filling slopes of contrast intensity profiles correlate well with manual frame counts and flow grades in both our derivation and validation datasets. Furthermore, the generated washout dynamics appear to provide different information to the traditional frame count and flow grade metrics, which only deal with the initial filling phase, suggesting that washout dynamics of the contrast intensity profiles may capture novel information about the coronary microcirculation.

## 1. Introduction

Coronary angiography enables visualization of the coronaries and is a foundation for the assessment of anatomic stenosis during heart catheterization. However, the ability to extract functional information on flow of contrast through the coronary tree has been lacking. Accurate and objective assessment of flow from coronary angiography has the potential to provide valuable prognostic information about the physiological relevance of stenoses, the adequacy of revascularization, as well as the microvascular function.

In clinical practice, flow is frequently described subjectively using the TIMI flow grading which involves expert evaluation of the bulk filling of the coronary tree.^1^ Alternatively, the TIMI frame count method was devised by Gibson et. al. to provide a more quantitative assessment of flow by counting the number of frames between dye entering the coronary ostium and arriving at pre-determined landmarks.^2–4^ This approach enables assessment of bulk flow but requires labor-intensive manual identification of the relevant time points. Other approaches to quantify flow through the coronary tree have been developed but typically require deployment of pressure sensitive catheters (FFR)^5,6^ or use of additional imaging modalities that provide largely estimates of flow derived from anatomical reconstructions (CT-FFR).^7,8^ Zhao et al previously leveraged a machine-learning segmentation strategy in an effort to quantify flow from angiograms but focused on correlation with FFR rather than on measures directly obtainable from diagnostic angiograms without additional imaging or catheterization time.^9^

Here we introduce an automated approach to quantitatively assess bulk flow from angiograms facilitated by the AngioNet neural network using the right coronary artery (RCA) as an example.^10^ We demonstrate that this approach provides quantification of flow that correlates well with TIMI frame counts without requiring manual identification of anatomical landmarks in angiograms. We also demonstrate unique features of this quantification that may provide additional insights beyond traditional measures, specifically with regards to microvascular function.

## 2. Methods

### I. Angiogram selection

A dataset of de-identified RCA angiograms of left anterior oblique (LAO) projections previously used as part of the AngioNet training cohort^10^ was the starting point of the current study. This dataset was acquired using a Siemens Artis Q Angiography system under study protocol (HUM00084689) approved by the Institutional Review Board of the University of Michigan Medical School (IRBMED). With low risk for subject harm and retrospective use of data collected for clinical purposes, IRBMED approved use without requiring informed consent.

Included patients were referred for a diagnostic coronary angiography procedure at the University of Michigan Hospital between 2018 and 2019. Patients with pacemakers, implantable defibrillators, chronic total occlusions, and prior bypass grafts were excluded. A subset of RCA angiograms was randomly selected from the full dataset with the additional inclusion criteria of having anatomy and angiographic technique allowing for TIMI frame counting. This subset was separated into two cohorts, consisting of 80 derivation and 197 validation videos.

### II. TIMI grade and TIMI frame count

Manual review of angiograms involved characterization of the TIMI grade (per the protocol described in Chesebro et. al., 1987) and corrected TIMI frame count (per the protocol originally published in Gibson et. al., 1996), hereafter called the TIMI frame count.^1,2^ Of note, TIMI frame counts were corrected for variations in acquisition frame rate and normalized to 15 frames per second. Angiograms were independently inspected by 3 reviewers in batches of 20 angiograms, with intervening mediation sessions to support consensus on TIMI frame count and grade between batches, until the Kendall Tau correlation coefficient for inter-rater reliability for the subsequent batch was >0.75. The raters then divided up and independently reviewed the remaining angiograms.

### III. Angiogram segmentation with AngioNet

De-identified angiograms were separated into individual PNG frames (Figure 1a). These frames were then recursively segmented by the AngioNet neural network^10^ (Figure 1b, and supplemental video). Briefly, AngioNet combines an Angiographic Processing Network (APN) coupled to a semantic segmentation network, Deeplabv3+.^11^ The APN underwent preliminary training to mimic several unsharp mask filters applied in series while the Deeplabv3+ network was initialized with pre-trained weights available from the TensorFlow Deeplab GitHub repository. The coupled network was subsequently trained on a combination of single angiogram frames obtained at the University of Michigan and the Madras Medical Mission, labeled by expert reviewers. Of note, the frames used for training were selected by identifying those with the maximum summed pixel intensity—corresponding to that with the most filled coronary tree. The network weights used for segmentation in the present work were the final weights from Iyer et al, 2021.^10^

**Figure 1.**
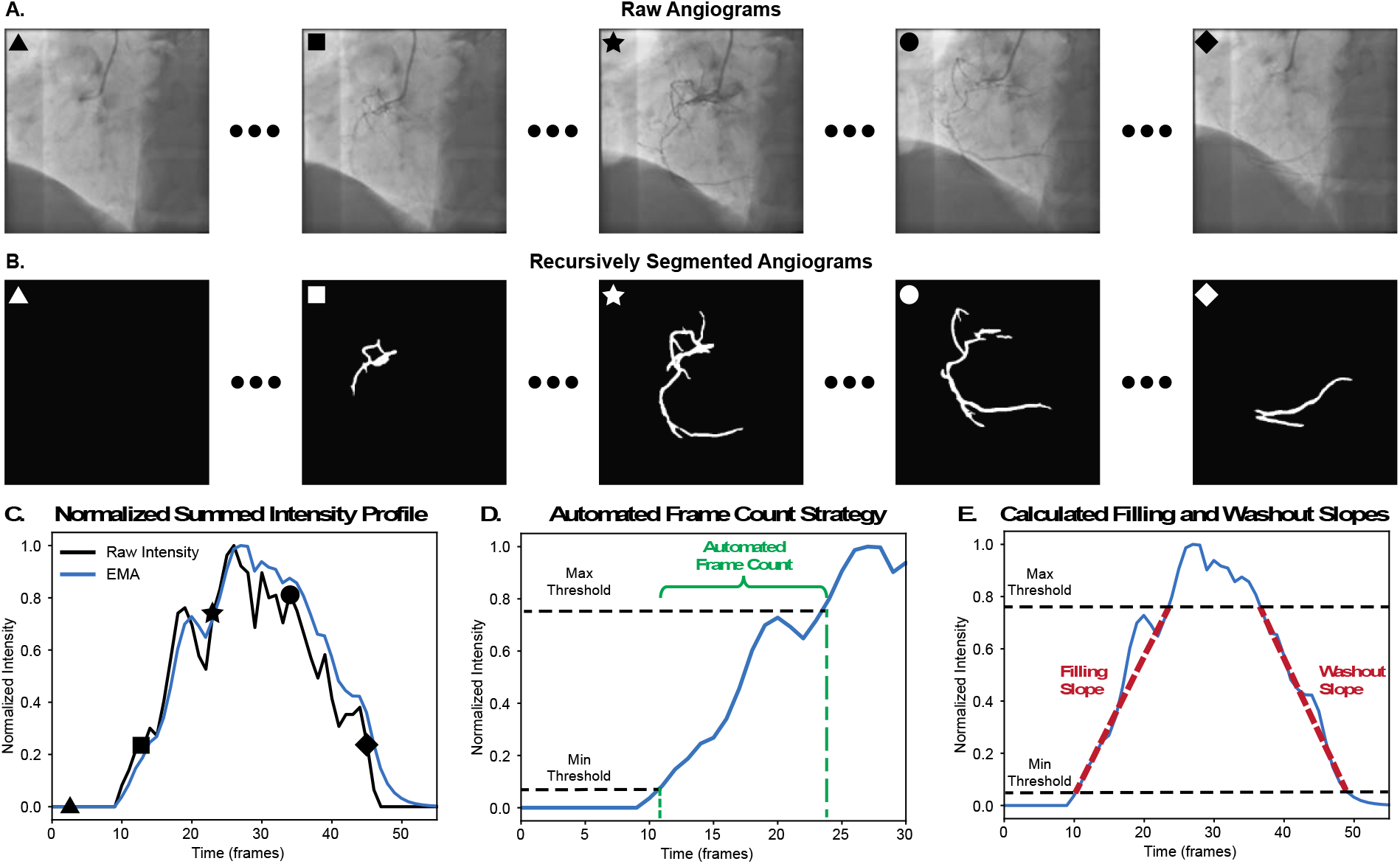
Procedure for generating contrast intensity profiles and determining automated frame count. **A**. Selected individual frames from an angiogram. **B**. Segmentations of corresponding frames using recursive application of AngioNet. **C**. Normalized summed intensity profile (black) generated by summing binary segmentation arrays from each frame with estimated moving average spline interpolation (blue). Markers indicate corresponding frames from above. **D**. Spline-smoothed intensity profile of filling phase with the beginning of filling (first crossing of minimum threshold intensity-bottom, black dashed line) and end of filling (first crossing of maximum threshold intensity-top, black, dashed line) frames indicated as green dashed lines. Automated frame count defined as number of frames between initial and final filling frames. **E**. Illustration of filling and washout slopes (red dashed lines) on full spline-smoothed intensity profile. The filling slope is defined as the mean slope of the intensity profile between the beginning of filling and end of filling, as defined in D. The washout slope is defined as that between the second crossing of the maximum threshold (beginning of washout) and second crossing of the minimum threshold (end of washout).

### IV. Optimization of Bulk flow assessment

Time series of the net intensity of angiograms were generated by summing the number of filled pixels in each frame (Figure 1c). These intensity time series were smoothed using an exponential moving window with smoothing factor of 0.4 and then normalized to the [0,1] range by subtracting the minimal intensity and dividing by the maximal intensity to produce the contrast intensity profiles leveraged for further analysis. B-spline basis functions were fit to individual nomograms to enable extrapolation of filling time points between individual frames at a resolution of 0.1 frames.

Based on these contrast intensity profiles, automated frame counts were calculated as depicted in Figure 1d. The derivation cohort was used to optimize parameters of the proposed automated frame count algorithm such that correlation with standard TIMI frame count was maximized while the validation cohort was subsequently used to test the performance of the optimized automated frame count. The initiation of filling was defined as the time when the spline profiles crossed a ‘minimal filling threshold’, defined as 0.05 (selected to avoid inclusion of segmentation artifacts, see discussion). The end of proximal filling was defined by a ‘maximal filling threshold’ set at 0.70 and empirically determined to optimize the correlation with the TIMI frame count in the derivation cohort (parameterization discussed in conclusions). The automated frame count was then defined as the number of frames between the minimal and maximal filling thresholds (potentially non-integer given spline fitting) and normalized for acquisition frame rate.

Following the initial algorithm optimization, the derivation and validation cohorts were pooled for analyses of filling and wash-out slopes. Using the same minimum and maximum filling thresholds as above, a filling slope was defined as the average rise in pixel intensity per frame between these thresholds (diagrammed in Figure1e). Identical maximum and minimum thresholds were applied in the reverse order to the washout slope of the intensity profile to enable calculation of mean washout slopes. Angiograms in which the minimum filling threshold was not reached during washout were omitted from the washout slope analysis. These generated slopes have units of normalized intensity/frame.

### V. Statistics

Analyses were performed within Python 3.9.13 with pandas, SciPy, and scikit-learn libraries. Continuous and discrete variables are reported as mean ± SD, [minimum – maximum]. Hypothesis testing leveraged Student’s t-tests for exploring differences in the mean of continuous data, given normality of data, and Kendall Tau correlation for assessing interrater reliability between discrete variables. A two-tailed observed z-test statistic was calculated to assess for a statistically significant difference between the correlations between TIMI frame counts and AngioNet-derived frame counts for the derivation and validation cohorts with a pre-specified *α* < 0.05.

## 3. Results

### I. Qualitative assessment of filling and washout

As previously quantitatively demonstrated by Iyer et al., the segmentation quality for individual angiogram frames is high. There is, however, substantial frame-to-frame variability in normalized intensity (see Figure 1c) which prompted the smoothing procedure. Figure 2a plots all the filling— phase contrast intensity profiles of the 80 angiograms in the derivation cohort and demonstrates the large variance in average slope between angiograms. There are also notable non-linearities within individual contrast intensity profiles (see discussion). Figure 2b plots all the washout intensity profiles for the derivation cohort. As with the filling portions of the profiles, there is large qualitative variance in the rate of dye washout and notable non-linearities within individual profiles (see discussion).

**Figure 2.**
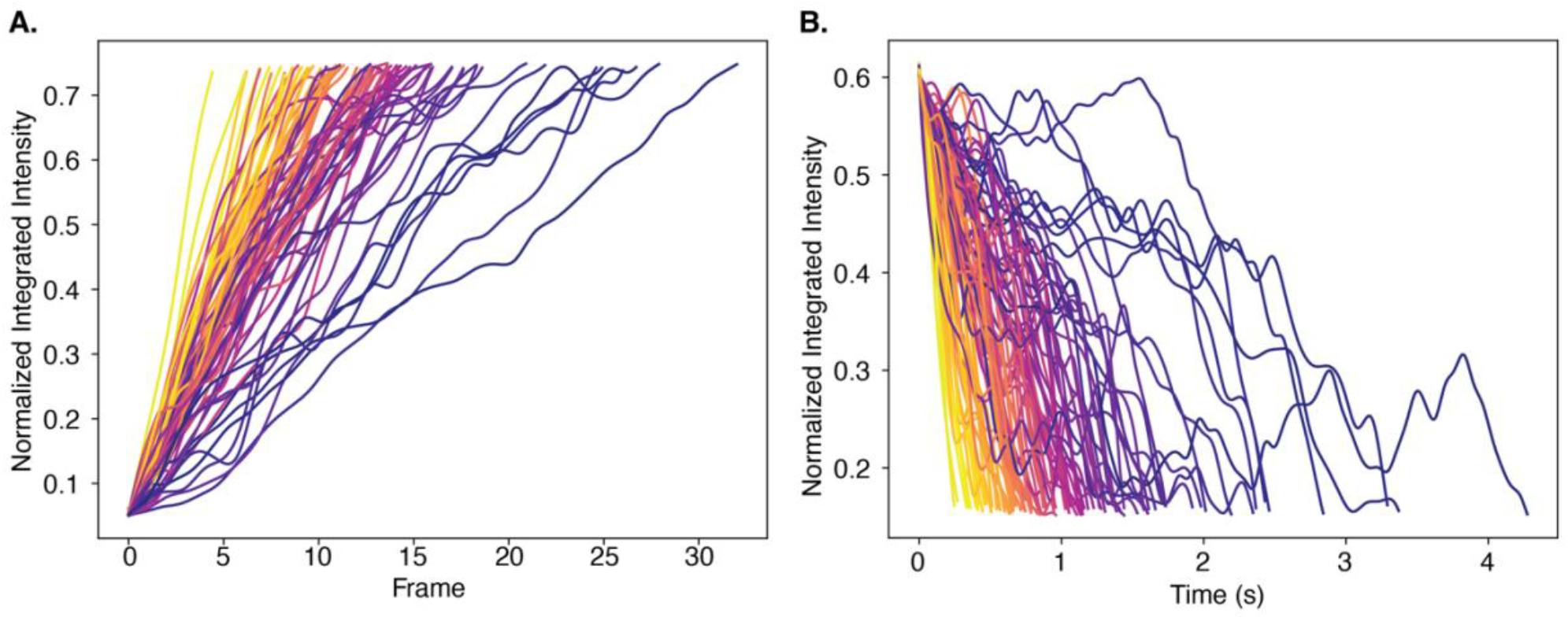
Heterogeneity of filling and washout intensity profiles. The filling. **A**. and washout **B**. portion of the normalized integrated intensity profiles are plotted against time for the 80 angiograms in the derivation cohort. Color-mapping is by mean slopes of profiles.

### II. Both derivation and validation cohorts exhibit strong correlations between TIMI and AngioNet-derived frame counts

Utilizing the 80 derivation angiograms, the segmentation flow count parameters (including smoothing function parameters, minimum and maximum filling thresholds) were manually adjusted to optimize the correlation between TIMI and AngioNet-derived frame counts. Figure 3a shows TIMI frame counts (13.6 ± 5.6 frames, [6 – 38 frames]) plotted on the abscissa against derivation dataset AngioNet-based frame counts (12.8 ± 4.5 frames, [5.7 – 29.7 frames]) plotted on the ordinate in blue circles. The trend line shows the least-squares regression fit with a R^2^ of 0.48 and t-test p << 0.001.

**Figure 3.**
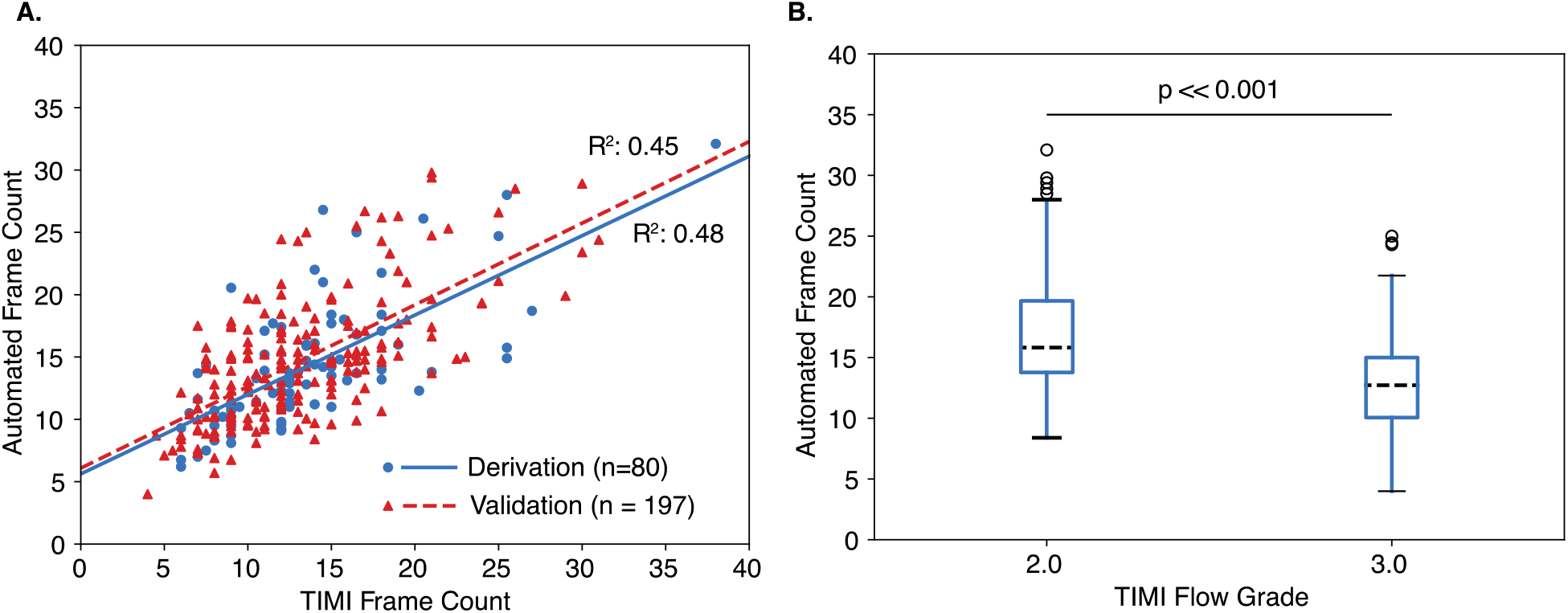
Automated frame counts are significantly correlated with TIMI frame counts and flow grades. **A**. TIMI frame counts are plotted on the abscissa against automated frame counts on the ordinate for the derivation (blue circles, n=80) and validation (red triangles, n=197) cohorts. Least squares correlations are plotted for derivation (solid blue line) and validation (red dashed line) cohorts with R^2^ of 0.48 and 0.45, respectively. **B**. Whisker plot distributions of automated frame counts of pooled cohorts plotted for TIMI flow grades of 2 and 3 with mean as black dashed line, blue box representing 25-75 percentile, and hashes marking 5^th^ and 95^th^ percentiles. Two-tailed T-test comparing population means with p << 0.001.

After this derivation process, the parameters were frozen, and the segmentation flow count process was applied to the validation cohort. The red triangle markers plotted in Figure 3a show TIMI frame counts (13.1 ± 5.1 frames, [4.0– 31 frames]) plotted against the validation dataset AngioNet-based frame counts (13.2 ± 4.6 frames, [3.8 – 28.4 frames]). The trend line shows the least-squares regression fit with a R^2^ of 0.45 and t-test p << 0.001. The observed z-test statistic comparing the correlation coefficients between the TIMI frame counts and AngioNet-derived frame counts for the derivation and validation cohorts was z = 0.7, corresponding to p = 0.48, thus failing to reject the null-hypothesis that the correlations are the same. Given the consistent performance across the two datasets, the angiograms were pooled for subsequent analyses.

Figure 3b shows boxplots of AngioNet-derived frame counts for the each TIMI flow grade included in the angiogram cohort. One angiogram with a flow grade of 1 was identified and excluded from analysis (a chronic total occlusion with right-to-right collaterals). The distributions of automated frame count for the angiograms with TIMI flow grade 2 (15.6 ± 5.0 frames, [7.8 – 29.7 frames]) and those with TIMI flow grade 3 (11.7 ± 3.6 frames, [3.8 – 23.2 frames]) were analyzed with a two-tailed t-test, yielding p << 0.001.

### III. Filling, but not washout, mean slopes correlate with TIMI frame counts

We next explored alternative parameters for characterizing bulk flow from the contrast intensity profiles by calculating the mean filling and washout slopes for each angiogram. We began by calculating the mean slope of the filling profile between the first crossings of the minimum and maximum thresholds (Figure 1e). Figure 4a shows TIMI frame counts plotted on the abscissa against mean filling slope on the ordinate (0.83 ± 0.29 1/s, [0.32 – 2.55 1/s]) for all pooled angiograms (n =277). The trend line in Figure 4a shows the least-squares regression fit with a R^2^ of 0.375 and corresponding t-test p << 0.001.

**Figure 4.**
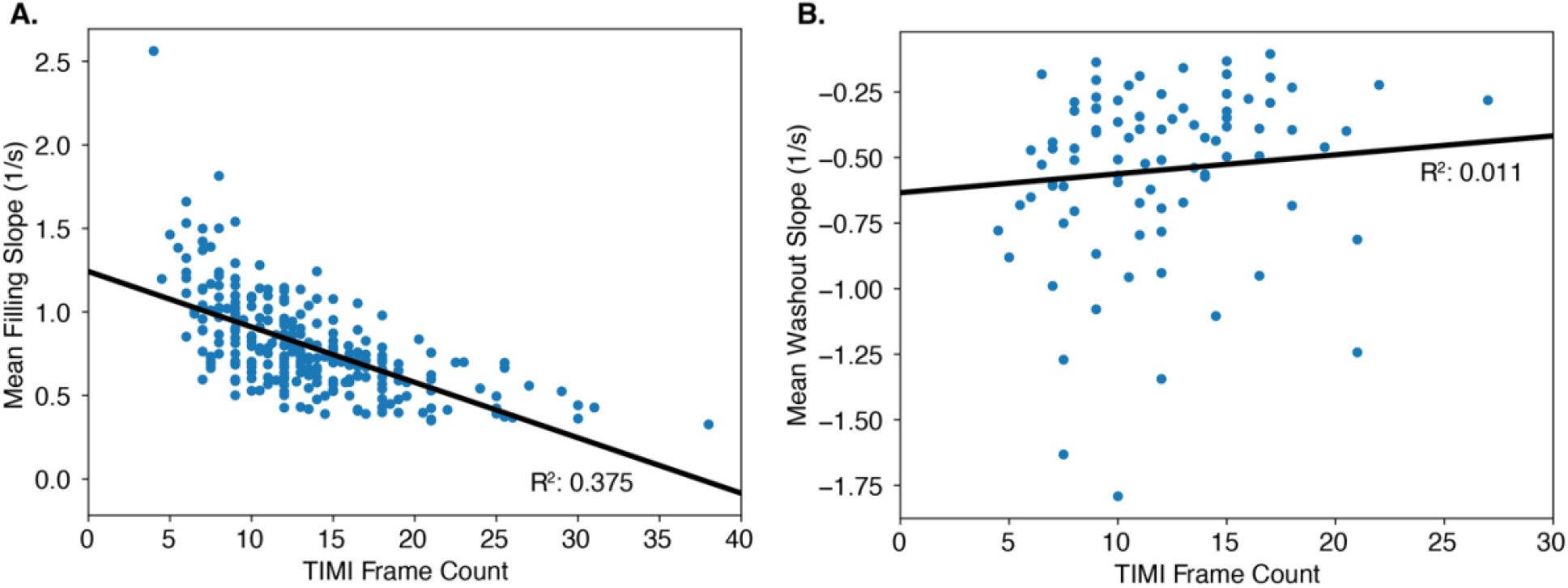
Mean filling, but not washout, slopes of contrast intensity profiles correlate with TIMI frame counts. **A.** Mean filling slope (in 1/s) plotted against TIMI frame count (n = 277). Black line is least squares regression with R^2^ of 0.375. **B**. Mean washout slope (in 1/s) plotted against TIMI frame count (n = 88). Black line is least squares regression with R^2^ of 0.011.

We next considered the washout portion of the intensity profiles. Only a subset of angiograms (n = 88) contained enough frames for the intensity profile to return below the minimum threshold. Figure 4b shows TIMI frame counts plotted on the abscissa against mean washout slope (−0.55 ± 0.33 1/s, [-1.79 – -0.11 1/s]) on the ordinate for this subset of angiograms. The trend line in Figure 4b shows the least-squares regression fit with a R^2^ of 0.011 and corresponding p-value of 0.33, indicating minimal association between TIMI frame counts and washout slope.

## 4. Conclusions

The current study demonstrates the feasibility of leveraging automated, machine-learning driven angiogram segmentation to perform quantitative assessment of bulk flow through coronary trees. This strategy involves segmentation of individual angiogram frames followed by automated quantification of the contrast in-flow frame count and mean slopes of both the filling and washout phases. While several parameters in the derivation process were tuned during the derivation procedure, the consistent performance of the technique within the validation cohort suggests it is generalizable (with the caveat of the exclusion criteria discussed below). Both the AngioNet-derived frame counts and mean filling slopes show significant, and relatively strong, correlations with TIMI frame counts. The performance of this correlation can likely be enhanced by using more sophisticated approaches to quantify contrast flow or velocity, and by improved segmentation quality, specifically geared to deal with dynamic data, rather than simply performing frame-by-frame segmentation. However, much of the residual variance between the AngioNet parameters and manual frame counts likely stems from the imperfection of the latter as a Gold Standard. One additional consideration is that segmentation frame counts may be more sensitive to the precise angle of acquisition, despite all angiograms being obtained from a LAO projection. An important next step in characterizing the performance of the proposed neural network-driven approach for flow quantification will be to assess its prognostic accuracy by exploring the potential correlation of parameters with cardiovascular outcomes.

The dynamics present in the contrast intensity profiles also point to the potential for generating richer insight into the flow dynamics within the coronary tree than have generally been accessible from simple diagnostic angiograms without machine-learning derived approaches. The contrast intensity profiles of individual angiograms exhibit frame-to-frame inflections that likely reflect technical differences such as dye injection technique (e.g., how well the catheter is engaged or the strength of injection) and physiologic variables such as injection phase relative to the cardiac cycle, though some portion of this variance may be due to artifactual discrepancies between segmentations of individual frames. Going forward, more sophisticated processing of segmented angiograms (i.e. mapping the centerline of filling coronaries onto co-registered cross-sectional imaging through deformable registrations) may enable more precise quantification of frame-by-frame variation in absolute flow velocities. Importantly, the AngioNet-derived mean washout slopes were notably not correlated with TIMI frame counts or flow grades. While variability in panning technique may underly some of the washout slope variability, we suspect the dynamics captured by the washout slopes may reflect the characteristics of the coronary microcirculation. Larger microvascular resistances may result in sluggish washout dynamics, which is not captured by current angiography interpretation techniques. Repeating this analysis in a cohort of angiograms with more consistent angiogram technique during the washout phase, and correlation with invasively determined index of microcirculatory resistance (IMR) will help elucidate this.^12^

### Parameter selection

The tunable parameters in the presented machine learning flow characterization approach are the strategy of contrast intensity profile smoothing and the minimum/maximum thresholds used. We trialed multiple different smoothing parameters and found that correlation with the manual counts was robust to the precise degree of smoothing but unsmoothed data led to poorer performance, likely due to the presence of frame-to-frame variability in segmentation accuracy. Improved segmentation (perhaps by training a neural network on full angiograms rather individual frames, using deformable registrations mapping vessels over the entire dynamic data) would potentially obviate the need for smoothing. Similarly, the minimal threshold was selected to mitigate the effect of small variations in segmentation prior to contrast injection. The maximum threshold was selected by manually tuning the parameter to optimal performance. Of note, this yielded a higher mean intensity than that at the final frame of the TIMI frame count strategy (when contrast first fills the first branch arising from the posterior lateral extension of the RCA after the origin of the posterior descending artery) for a subset of 10 angiograms, likely due to the delay imposed by the smoothing approach

## Limitations

In this proof-of-concept study, we have focused on the RCA given the simpler anatomy of the right coronary tree. Generalization of this approach to the left coronary tree will benefit from further segmentation of angiograms to label individual branches of the LCA. The presented technique for quantifying filling is also limited by its reliance on the dimensionless quantity of number of pixels labeled as vessel within the entire RCA. Expanding the segmentation ability of AngioNet to label individual branches could enable this technique to be applied to individual coronary vessels, potentially identifying flow-limitations on a vessel-by-vessel basis. Finally, utilization of a more advanced algorithm for tracking flow, like center line tracking, could enable quantification of flow and velocity in individual vessels.

## Summary

Even with the above limitations, the current technique compares promisingly with the currently employed methods for flow quantification from diagnostic angiograms. It provides a more rigorous and standardized method to assess dye filling while also rendering novel information on the differences in washout dynamics. To directly assess the clinical utility of these techniques, future research is needed to determine how well the AngioNet-derived flow quantities correlate with subsequent cardiovascular clinical outcomes.

## Data Availability

All data produced in the present study are available upon reasonable request to the authors. The trained weights of the AngioNet Neural Network are proprietary property of AngioInsight and cannot be shared.

## 6. Acknowledgments

Kritika Iyer and co-authors of the AngioNet neural network.

